# Misclassification of heritable mortality undermines estimates of intrinsic life span heritability

**DOI:** 10.64898/2026.02.26.26347172

**Authors:** Fergus Hamilton

## Abstract

In a recent article in *Science*, Shenhar et al. report that human life span heritability reaches ∼55% after removing “extrinsic” mortality, roughly seven-fold higher than recent large pedigree estimates. This conclusion rests on classifying deaths from infections and accidents as environmental noise independent of genetics. This premise is biologically untenable: susceptibility to severe infection is substantially heritable, with adoptee studies showing relative risks exceeding 5 for infection death when a biological parent died of infection. By encoding the assumption that extrinsic mortality is non-genetic directly into their Gompertz-Makeham model, removing it necessarily inflates heritability estimates. This creates selection bias rather than correcting for confounding and explains the contradiction with both pedigree studies and GWAS findings. The proposed heritability estimate is therefore not the true heritability of any population, past or present.

---

In a recent article in *Science*, Shenhar et al. propose that lifespan heritability is approximately 55% after correcting for “extrinsic mortality” - deaths from “external factors such as accidents… [and] infectious diseases.”(*1*) Their core premise is that extrinsic mortality represents purely environmental noise that masks genetic signal. This premise is biologically untenable, and the methodology introduces bias rather than removing it.

## Infection mortality is highly heritable

Susceptibility to severe infection is profoundly heritable, supported by decades of immunogenetic research. The HLA/MHC region is among the most polymorphic in the human genome, consistent with long-term balancing selection that includes pathogen-mediated pressures.(*2*) Human blood cell variation is a major driver of susceptibility to malaria, a leading cause of death in children for thousands of years.(*3*–*8*)

This heritability persists into later life. Sørensen et al, studying 960 Danish adoptees, found that death of a biological parent from infection before age 50 conferred a relative risk of 5.81 (95% CI: 2.47–13.7) for infection-related death in the adoptee - substantially higher than for cardiovascular causes (RR 4.52) or cancer (RR 1.19). Crucially, death of an adoptive parent from infection conferred no increased risk (RR 0.73, 95% CI: 0.10–5.36).(*9*) Danish twin studies subsequently confirmed heritability of infectious disease mortality at approximately 40%, with monozygotic concordance significantly exceeding dizygotic concordance.(*10*)

Because infections are included among ‘extrinsic’ causes, treating extrinsic mortality as a component that is fixed across individuals risks excluding genetically mediated variation operating through immune susceptibility.

### The model encodes the assumption

Shenhar et al. decompose mortality into extrinsic and intrinsic components:

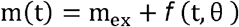

where m_ex_ is extrinsic mortality and θ represents intrinsic ageing parameters. In their framework, genetic variation operates through θ, while m_ex_ is treated as a cohort-level constant shared across individuals. They define “heritability of intrinsic lifespan” (HIL) by extrapolating to a counterfactual world where m_ex_ = 0.

This formulation embeds the assumption that genetic variation does not influence extrinsic mortality. Under this specification, setting m_ex_ to zero necessarily increases the apparent genetic contribution to remaining mortality - the conclusion follows directly from the model structure. If genetics genuinely affects susceptibility to causes classified as “extrinsic” (such as infection), this assumption is violated, and the ∼55% figure describes a counterfactual population, not a correction for confounding.

### Selection bias, not confounding correction

Removing extrinsic mortality, whether by excluding early deaths or extrapolating to m_ex_ = 0, conditions on survival. If genetic factors influence both extrinsic death risk and later-life mortality, survival is a collider, and conditioning on it induces bias.

This shared genetic influence is biologically highly plausible. Genetic variants affecting infection susceptibility or stress resilience operate through immune function, inflammation, and cellular repair the same pathways driving “intrinsic” ageing. The assumption that genetic effects on extrinsic and intrinsic mortality are independent contradicts the pleiotropic nature of genes influencing frailty.

Simulation (**Supplement S1**) confirms this: when genetics affects both extrinsic death risk and intrinsic lifespan, Falconer’s h^2^ increases after removing extrinsic deaths - replicating Shenhar et al.’s pattern - even with no change to underlying genetic architecture. Rising heritability after “correction” cannot distinguish noise removal from selection bias. The adoption literature, showing strong familial clustering of infection mortality through biological but not adoptive parents, favours selection.

### Inconsistent treatment across the lifespan

The restriction to young-middle age is also inconsistent. National mortality data show a U-shaped relationship between age and non-infectious extrinsic mortality - falls, motor vehicle accidents, and similar causes increase substantially at older ages.(*11*) If extrinsic mortality warrants removal at age 30, it should equally warrant removal at age 80. The selective application of this “correction” only at younger ages is methodologically incoherent.

### The additivity assumption ignores gene-environment interaction

The Gompertz-Makeham framework assumes intrinsic and extrinsic mortality are independent and additive on the hazard scale. This violates the biological reality of gene-environment interaction. Genetic risk factors frequently manifest through susceptibility to environmental insults - the sickle cell trait’s phenotypic effect depends on malaria exposure.(*12*) By assuming additivity, Shenhar et al. treat gene-environment interactions as pure environmental noise to be removed, when these represent a primary mechanism by which genes influence mortality.

### Infection remains relevant - historically and today

One might argue that infection mortality is a historical phenomenon irrelevant to modern populations. This defence fails on multiple grounds. First, the cohorts Shenhar et al. analyse are themselves historical - Danish twins born 1870–1900, Swedish twins born 1886–1925, SATSA twins born 1900–1935. These are precisely the populations where infectious mortality was highest.

Second, the premise that infection no longer matters is empirically false. Pneumonia and influenza consistently rank among the top 10 causes of death in high-income countries.(*13*) COVID-19 demonstrated - complete with GWAS identifying severity loci at *OAS1, TYK2*, and elsewhere - that infection susceptibility remains genetically influenced and consequential in contemporary populations.(*14*)

Finally, the distinction between infectious and “intrinsic” mortality is not clean. Immune function is not only critical for pathogen defence but also drives inflammation, atherosclerosis, and cancer surveillance. By classifying infection deaths as non-genetic noise, the authors erroneously discard the genetic contribution of the immune system to the “intrinsic” aging processes they claim to isolate.

### Contradiction with other evidence

The resulting estimate of ∼55% contradicts both pedigree and molecular evidence. Ruby et al. (2018), using millions of genealogical records with correction for assortative mating, estimated life span heritability below 7%.(*15*) Shenhar et al. dismiss this as reflecting “environmental heterogeneity,” but this reasoning is circular: they assume their additive model is correct to explain away data that contradicts it.

The GWAS literature provides independent evidence against high heritability. If lifespan were truly 50% heritable, we would expect genome-wide association studies to have identified substantial explained variance. Instead, the largest longevity GWAS identify only a handful of robust loci - *APOE, FOXO3*, and few others - collectively explaining a small fraction of variance.(*16*)

## Conclusion

The approach used by Shenhar et al. cannot support the claim that lifespan heritability approaches 50%. Their method of removing extrinsic mortality relies on an assumption contradicted by immunogenetic evidence and introduces selection bias rather than correcting for confounding. The ∼55% figure applies only to a selected, biologically implausible counterfactual population-not to any actual human population past or present.

## Supporting information

Sim.R

## Data Availability

All data is included with the study.

## Notes

### Competing Interest Statement

The authors have declared no competing interest.

### Funding Statement

This study did not receive any funding.

