## Supplement S1 for "Misclassification of heritable mortality undermines estimates of intrinsic life span heritability"

**Methods**

We simulated 50,000 monozygotic (MZ) and 50,000 dizygotic (DZ) twin pairs. For MZ pairs, both twins share identical genetic values (G); for DZ pairs, genetic values are correlated at 0.5. Each twin's frailty was modelled as a weighted sum of genetic and independent environmental components. Extrinsic death probability was generated as a logistic function of frailty, so that higher frailty increased the probability of early extrinsic death. Lifespan among survivors was modelled as a linear function of frailty plus noise; those who died extrinsically were assigned an early death age.

We computed Falconer's heritability estimate, h² = 2(r_MZ_ − r_DZ_), in (i) the full cohort and (ii) the subset of pairs where both twins survived without extrinsic death. The genetic weight parameter was calibrated so that the "corrected" heritability approximated 0.45, similar to Shenhar et al.'s estimates.

This setup isolates the key mechanism: when extrinsic death risk is genetically influenced, conditioning on survival induces selection bias. Removing extrinsic deaths enriches the sample for lower-frailty genotypes and increases twin correlations and therefore Falconer h², even without any change to the underlying genetic architecture. A rise in heritability after removing "extrinsic" deaths does not by itself demonstrate that environmental noise was removed; it is equally consistent with selection on genetically-patterned mortality.

Shenhar et al. argue their findings are robust because a sensitivity analysis - modelling extrinsic mortality as the sum of a constant and an exponentially rising term yielded identical heritability estimates. Their sensitivity analysis constrains the exponential slope of extrinsic mortality to 80% of intrinsic mortality. Because intrinsic mortality rises faster with age, this constraint mathematically ensures that at older ages (where most deaths occur in the twin cohorts analysed), nearly all deaths are classified as 'intrinsic'. The 'robustness check' therefore changes almost nothing about the age range where twin correlations are estimated, rendering it uninformative about whether extrinsic mortality is genetically influenced

**Figure 1:** Effects on heritability driven by selection. A: Genetic frailty (G) distribution for all twins, those with any extrinsic death, and those where both twins survive. Removing extrinsic deaths enriches for lower G. B: Falconer h² estimated from twin correlations rises after excluding extrinsic deaths, even though the only change is selection on survival. This is a toy example to show the effect of selection bias and clearly the degree of bias reflects the input parameters of the model.

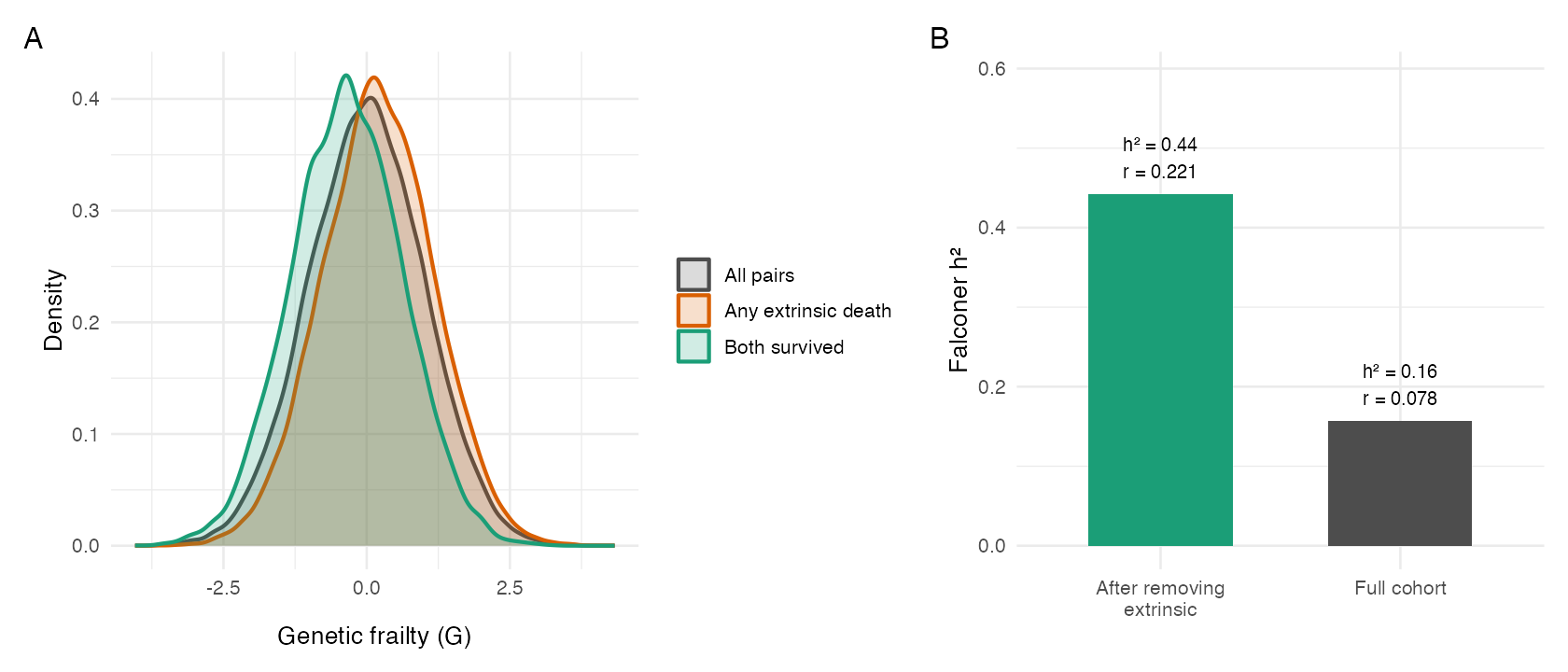

Code:

```
#!/usr/bin/env Rscript
# Selection-bias simulation with MZ + DZ twins and Falconer h²

set.seed(42)

if (!requireNamespace("ggplot2", quietly = TRUE)) {
  stop("Package 'ggplot2' is required for plotting.")
}
if (!requireNamespace("patchwork", quietly = TRUE)) {
  stop("Package 'patchwork' is required for plotting.")
}
library(ggplot2)
library(patchwork)

n_pairs <- 50000

# Genetic and environmental components
g_weight <- 0.74
e_weight <- 0.61
target_h2_corr <- 0.45

# Extrinsic risk (logistic on frailty)
extrinsic_slope <- 0.62
target_extrinsic <- 0.45
extrinsic_shift <- 0.70

logistic <- function(x) 1 / (1 + exp(-x))

simulate_twins <- function(n, r_g, u_ex1, u_ex2, eps1, eps2, u_ed1, u_ed2, g_weight) {
  # Genetic correlation r_g: 1.0 for MZ, 0.5 for DZ
  g1 <- rnorm(n)
  g2 <- r_g * g1 + sqrt(1 - r_g^2) * rnorm(n)

  e1 <- rnorm(n)
  e2 <- rnorm(n)

  frailty1 <- g_weight * g1 + e_weight * e1
  frailty2 <- g_weight * g2 + e_weight * e2

  p_ex1 <- logistic(extrinsic_slope * (frailty1 - extrinsic_shift))
  p_ex2 <- logistic(extrinsic_slope * (frailty2 - extrinsic_shift))
  d1 <- u_ex1 < p_ex1
  d2 <- u_ex2 < p_ex2

  L1 <- 80 - 5 * frailty1 + eps1
  L2 <- 80 - 5 * frailty2 + eps2
  if (sum(d1) > 0) L1[d1] <- 30 + u_ed1[1:sum(d1)]
  if (sum(d2) > 0) L2[d2] <- 30 + u_ed2[1:sum(d2)]

  list(
    L1 = L1,
    L2 = L2,
    extr1 = d1,
    extr2 = d2,
    g1 = g1,
    g2 = g2
  )
}

falconer_h2 <- function(mz, dz) {
  r_mz <- cor(mz$L1, mz$L2)
  r_dz <- cor(dz$L1, dz$L2)
  list(r_mz = r_mz, r_dz = r_dz, h2 = 2 * (r_mz - r_dz))
}

# Pre-draw noise for comparability
set.seed(101)
u_ex1 <- runif(n_pairs)
u_ex2 <- runif(n_pairs)
eps1 <- rnorm(n_pairs, 0, 5.8)
eps2 <- rnorm(n_pairs, 0, 5.8)
u_ed1 <- rnorm(n_pairs, 0, 4.8)
u_ed2 <- rnorm(n_pairs, 0, 4.8)

# Calibrate g_weight so corrected h² ~ target_h2_corr
calibrate_g <- function(target, lo = 0.1, hi = 2.0) {
  for (i in 1:40) {
    mid <- (lo + hi) / 2
    mz_tmp <- simulate_twins(n_pairs, 1.0, u_ex1, u_ex2, eps1, eps2, u_ed1, u_ed2, mid)
    dz_tmp <- simulate_twins(n_pairs, 0.5, u_ex1, u_ex2, eps1, eps2, u_ed1, u_ed2, mid)
    keep_mz <- !mz_tmp$extr1 & !mz_tmp$extr2
    keep_dz <- !dz_tmp$extr1 & !dz_tmp$extr2
    h2_corr <- falconer_h2(
      list(L1 = mz_tmp$L1[keep_mz], L2 = mz_tmp$L2[keep_mz]),
      list(L1 = dz_tmp$L1[keep_dz], L2 = dz_tmp$L2[keep_dz])
    )$h2
    if (h2_corr < target) {
      lo <- mid
    } else {
      hi <- mid
    }
  }
  (lo + hi) / 2
}

g_weight <- calibrate_g(target_h2_corr)

mz <- simulate_twins(n_pairs, 1.0, u_ex1, u_ex2, eps1, eps2, u_ed1, u_ed2, g_weight)
dz <- simulate_twins(n_pairs, 0.5, u_ex1, u_ex2, eps1, eps2, u_ed1, u_ed2, g_weight)

# Extrinsic death fraction (individuals)
extr_frac_mz <- mean(c(mz$extr1, mz$extr2))
extr_frac_dz <- mean(c(dz$extr1, dz$extr2))

# Full sample
full <- falconer_h2(mz, dz)

# Corrected sample: remove extrinsic deaths in either twin
keep_mz <- !mz$extr1 & !mz$extr2
keep_dz <- !dz$extr1 & !dz$extr2

mz_corr <- list(L1 = mz$L1[keep_mz], L2 = mz$L2[keep_mz])
dz_corr <- list(L1 = dz$L1[keep_dz], L2 = dz$L2[keep_dz])
corr <- falconer_h2(mz_corr, dz_corr)

cat(strrep("=", 65), "\n", sep = "")
cat("SELECTION BIAS WITH MZ + DZ TWINS (Falconer h²)\n")
cat(strrep("=", 65), "\n\n", sep = "")
cat(sprintf("Pairs: %d\n", n_pairs))
cat(sprintf("Calibrated g_weight: %.3f (target corrected h² ≈ %.2f)\n", g_weight, target_h2_corr))
cat(sprintf("Extrinsic deaths (individuals): MZ=%.1f%%, DZ=%.1f%%\n\n",
            100 * extr_frac_mz, 100 * extr_frac_dz))
cat(sprintf("MZ kept: %d (%.1f%%)\n", sum(keep_mz), 100 * mean(keep_mz)))
cat(sprintf("DZ kept: %d (%.1f%%)\n\n", sum(keep_dz), 100 * mean(keep_dz)))
cat("Full sample:\n")
cat(sprintf("  r_MZ = %.3f, r_DZ = %.3f, h² = %.3f\n", full$r_mz, full$r_dz, full$h2))
cat("Corrected (remove extrinsic):\n")
cat(sprintf("  r_MZ = %.3f, r_DZ = %.3f, h² = %.3f\n\n", corr$r_mz, corr$r_dz, corr$h2))

# Plot: Falconer h² (full vs corrected)
h2_df <- data.frame(
  group = c("Full cohort", "Corrected\n(remove extrinsic)"),
  h2 = c(full$h2, corr$h2),
  r_mz = c(full$r_mz, corr$r_mz),
  r_dz = c(full$r_dz, corr$r_dz)
)
h2_df$label <- sprintf("h²=%.2f\nr_MZ=%.2f\nr_DZ=%.2f", h2_df$h2, h2_df$r_mz, h2_df$r_dz)

p_h2 <- ggplot(h2_df, aes(x = group, y = h2, fill = group)) +
  geom_col(width = 0.6) +
  geom_text(aes(label = label), vjust = -0.4, size = 3.2) +
  scale_fill_manual(values = c("Full cohort" = "#4D4D4D", "Corrected\n(remove extrinsic)" = "#1B9E77")) +
  scale_y_continuous(limits = c(0, min(1, max(h2_df$h2) + 0.2))) +
  labs(x = NULL, y = "Falconer h²", fill = NULL) +
  theme_minimal(base_size = 12) +
  theme(legend.position = "none")

# Density plot (use MZ for illustration)
g_all <- c(mz$g1, mz$g2)
g_extr <- c(mz$g1[mz$extr1], mz$g2[mz$extr2])
g_surv <- c(mz$g1[!mz$extr1], mz$g2[!mz$extr2])

dens_df <- data.frame(
  G = c(g_all, g_extr, g_surv),
  group = c(
    rep("All individuals", length(g_all)),
    rep("Any extrinsic death", length(g_extr)),
    rep("Survivors only", length(g_surv))
  )
)

p_den <- ggplot(dens_df, aes(x = G, color = group, fill = group)) +
  geom_density(alpha = 0.2, linewidth = 0.9) +
  scale_color_manual(values = c("All individuals" = "#4D4D4D", "Any extrinsic death" = "#D95F02", "Survivors only" = "#1B9E77")) +
  scale_fill_manual(values = c("All individuals" = "#4D4D4D", "Any extrinsic death" = "#D95F02", "Survivors only" = "#1B9E77")) +
  labs(x = "Genetic frailty (G)", y = "Density", color = NULL, fill = NULL) +
  theme_minimal(base_size = 12) +
  theme(legend.position = "right")

combo <- (p_den + p_h2) + plot_annotation(tag_levels = "A")

ggsave("fig_selection_bias_h2_dz.png", p_h2, width = 6.5, height = 4.2, dpi = 160)
ggsave("fig_selection_bias_density_dz.png", p_den, width = 7, height = 4.5, dpi = 160)
ggsave("fig_selection_bias_combined_dz.png", combo, width = 10.5, height = 4.5, dpi = 160)
```
